# Malaria; risk factors within urban and rural settings in the Sahelian region of Cameroon and the role of insecticide resistance in mosquitoes

**DOI:** 10.1101/2022.09.14.22279962

**Authors:** Samuel Fru Ngwa, Raymond Babila Nyasa, Seraphine Nkie Esemu, Vincent P. K. Titanji

**Author notes:** **Corresponding author:** Raymond Babila Nyasa, Department of Microbiology and Parasitology, University of Buea, P. O. Box 63, Buea, South West Region, Cameroon.

## Abstract

**Background:** Cameroon is among the 11 countries that account for 92 % of malaria infection in sub-Saharan-Africa in 2018, and Maroua III Health District and her environs witnessed a malaria outbreak in 2013 with hundreds of deaths. This study was aimed at understanding the risk factors of malaria in the urban and rural population and to investigate the level of mosquito’s resistance to Deltamethrin and Permethrin.

**Methods:** It was a cross-sectional community-based study carried out from August to October 2019, in which questionnaires were administered to 500 participants, to obtain information on demographics, socioeconomic, behavioral, and environmental factors thought to be associated with malaria infection in both rural and urban settings. Blood samples were collected for the diagnosis of malaria. Logistic regression analysis was used to identify risk factors for malaria. Mosquito resistance to Deltamethrin and Permethrin were investigated using the CDC Bottle Bioassay test.

**Results:** Malaria prevalence was 52.2%, which was significantly higher (p = 0.016) in rural areas (57.6%) as compared to urban areas (46.8%). The overall prevalence of asymptomatic malaria parasitemia in the population was 43.4% in contrast to symptomatic malaria which was 8.8%. In rural areas, risk factors were households using treated bed nets older than 3 years (AOR: 2.45 95% CI: 1.30 to 4.61 P-value: 0.006); households whose water source are unprotected wells (AOR: 3.04 95% CI: 1.21 to 7.64 P-value: 0.018). In urban areas, risk factors were households surrounded by farmland with crops (AOR: 2.08 95% CI: 1.14 to 3.80 P-value 0.017) and households using treated bed nets older than 3 years (AOR: 2.70 95% CI: 1.52 to 4.78 P-value 0.001). The age group 2-10years was significantly associated (p<0.001) with malaria in both rural and urban settings of the district. The geometric mean parasite density was found to decrease with increasing age of participants in the entire health district. The overall mortality of *Anopheles species* was 93.57% (91.19% in rural and 95.83% in urban areas) for deltamethrin, which was more sensitive than 83.85% (85.24% in rural and 82.46% in urban areas) for permethrin.

**Conclusion:** Relevant data for malaria control in Maroua III health district, a typical Sahelian environment, has been generated, and indicates that most of the burden of malaria is borne by school children. Deltamethrine was more effective than permethrine in the control of mosquito populations within these areas.

## BACKGROUND

Malaria remains one of the major public health problems in Africa and in 2019, the WHO African Region accounted for 94% of malaria cases globally [1] while in Cameroon all of its inhabitants live in malaria endemic areas with 71% living in high transmission areas [2], [3]. Cameroon is one of the 15 countries that accounts for nearly 80 % of malaria deaths globally [4] and this infection is endemic in Cameroon, with the degree of prevalence varying from one ecological zone to another [5]. The proportion of deaths due to malaria is highest in the Northern Regions (26% in the Far North and 27% in the North Region) where the malaria season is shortest [2]. In 2013, the Far North Region of Cameroon witnessed an upsurge in malaria infection, where more than 10,000 people were treated for malaria, within a period of one month in Maroua town alone, and more than 600 people lost their lives to malaria, within that period [6]. In unpublished data from the Far North Regional Delegation of Public Health which covers Maroua III health district, 51776 cases of malaria were recorded in the first quarter of 2019, with an infant mortality rate of 37.25% [7]. The current study is aimed at determining the prevalence, risk factors and role of insecticide resistance in Maroua III health district.

Maroua III health district comprises urban and rural settlements and is part of the Sahel region of Cameroon with a hot semi-arid climate and a lowland topography with poor drainage pattern, causing stagnant water in most neighborhoods during the raining season. The presence of standing water around habitats is also caused by human activities, wherein those in rural areas empty their waste water in surrounding bushes around their residents while those in urban areas empty their waste water from their kitchens and bathrooms onto the road, which creates sites for mosquito breeding. Rural areas are poorly constructed with thatched houses, grass roofs and earth floors, which allows mosquito movement in and out of the building, as opposed to urban areas where houses are better constructed with cement blocks, aluminum roofing sheets and cemented floors. Also, most neighborhoods in rural areas of the district are surrounded by bushes and farmland which can serve as mosquitoes’ habitat, unlike in urban areas where bushy environments and farmland are not common, but inhabitants live in crowded neighborhoods. However, subsistence animal husbandry is common practice in both rural and urban settings and in most cases these animals (goats, sheep, cattle) live with their owners in the same house and could serve as an alternative source of blood meal for mosquitoes. It is therefore necessary to investigate if these factors are associated with the risk of malaria infection in this health district.

Since 2000, progress in malaria control has resulted primarily from expanded access to vector control interventions particularly in sub-Saharan Africa where long-lasting insecticidal nets (LLINs) usage is the mainstay of malaria prevention strategies and in Cameroon 50% of the population had access to LLINs [8]. Mass distribution of LLINs throughout the country was implemented in 2011, with the distribution of approximately 8,654,731 LLINs [9], followed by a second round in 2015 and a third round took place in 2019 in which eight million LLINs were distributed in the national territory, including the Far North Region [1], in the same year, when this study was carried out. The commonly used preventive measures against malaria in the Maroua III Health District include; usage of LLINs, screening windows of buildings with nets and indoor residual spray with insecticides, coupled with intermittent preventive treatment of pregnant women and prophylactic treatment of children 6-59 months old. The geographic expansion of insecticide resistance in female *Anopheles* mosquitoes could be due to the fact that many countries do not carry out adequate routine monitoring for insecticide resistance in local vectors and monitoring data are often not reported in a timely manner [10]. The WHO Global report on insecticide resistance in malaria vectors from 2010–2016 also showed that resistance to pyrethroids, the commonly used insecticide class for LLINs, is widespread in all major malaria vectors across the WHO regions of Africa, the Americas, South-East Asia, the Eastern Mediterranean and the Western Pacific [10]. Despite the usage of these LLINs in Maroua III health district, inhabitants still complain of mosquito bites when their bodies are in contact with the bed net at night. This could be due to mosquito resistance to pyrethroids used in treated bed nets or LLINs, which permit the mosquito to land on the net and bite its occupants through the pores of the bed nets using its proboscis. It is therefore important to determine if mosquitoes in rural and urban settings in Maroua III health district are susceptible/resistant to the commonly-used pyrethriods; Delthametrin and permethrin.

## METHODS

### Study Area

Maroua is the capital of the Far North Region of Cameroon, located at longitude 14.3210° E and latitude 10.5925° N. This study was conducted in localities within Maroua III health district, which consists of rural and urban settlements and is divided into ten (10) health areas (Fig 1) having rural and urban settlements. This area is part of the Sahel region of Cameroon with a hot climate and is characterized by strong winds during the short raining season that lasts from June to October and high temperatures during the dry season in the months of November to May. The area is occupied mostly by Maroua city dwellers who work within the city and peasant farmers in the various villages, who are mostly Fulani Muslims. The common language spoken in both urban and rural areas is Fulfulde.

**Fig 1:**
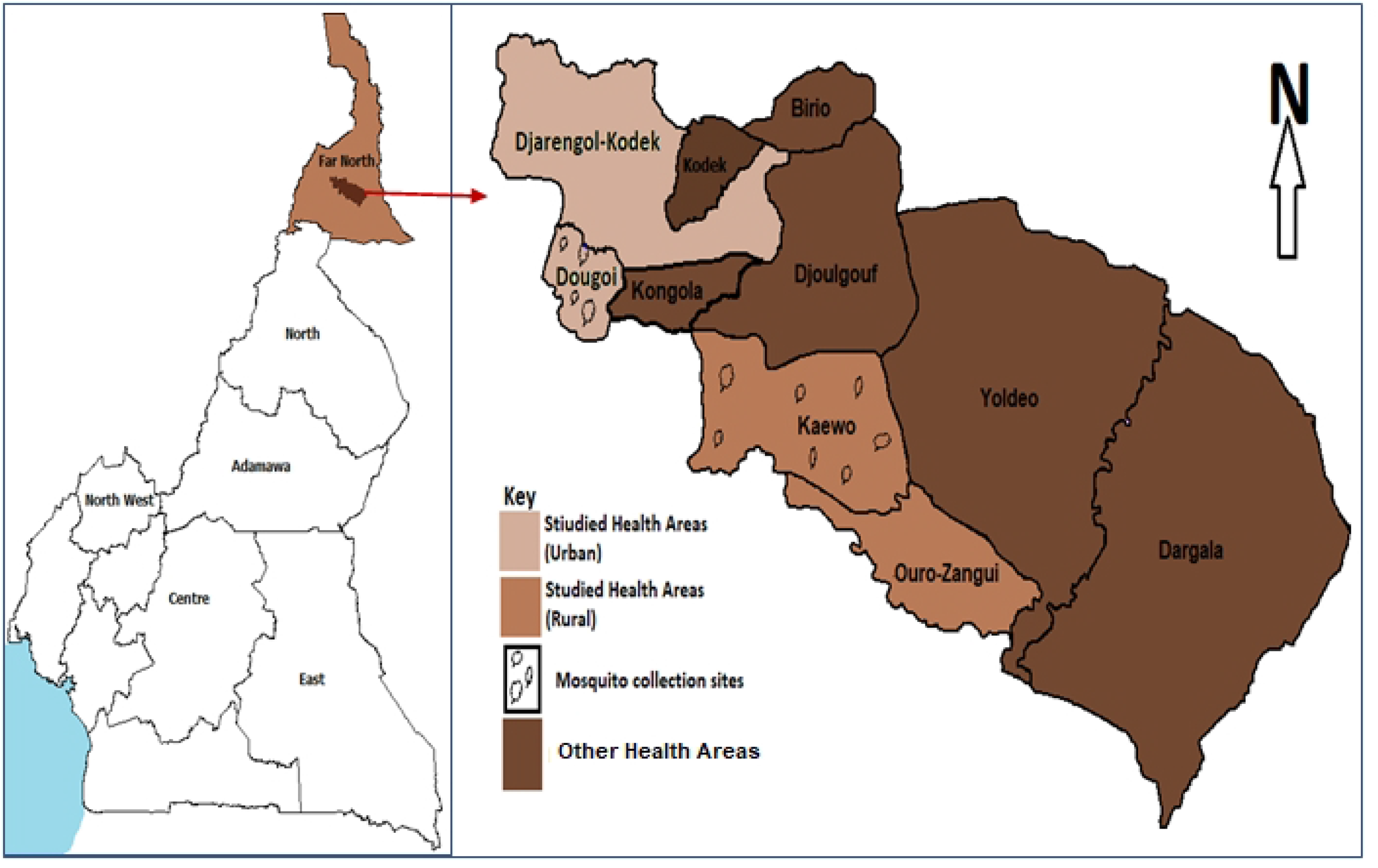
Map of Maroua III health district showing the various health areas where data was obtained from participants and mosquitoes collection sites. *Source: Rasta map from MinSante (Cameroon Ministry of Public Health, www.minsante.cm) and redrawn to scale by research team*

### Study Design

A cross-sectional study was carried out involving in-depth interviews of participants from house to house in both rural and urban communities of Maruoa III health district using a structured questionnaire from August to September 2019. Pretesting of the questionnaire was carried out in Meskine health area, which is also found in the Far North Region. Two health areas, Kaewo and Ouro-Zangui were randomly selected from balloting, using the names of eight health areas on folded and twisted pieces of paper for sample collection and questionnaire administration in rural settlements. Meanwhile, the two urban areas, Djarengol-kodek and Dougoi, were included in the study for sample collection and questionnaire administration for urban settlements as shown in Fig 1. The Study profile for households and individuals selected in both rural and urban areas is illustrated in Fig 2, whereby some households were vacant, and some households refused to participate.

**Fig 2:**
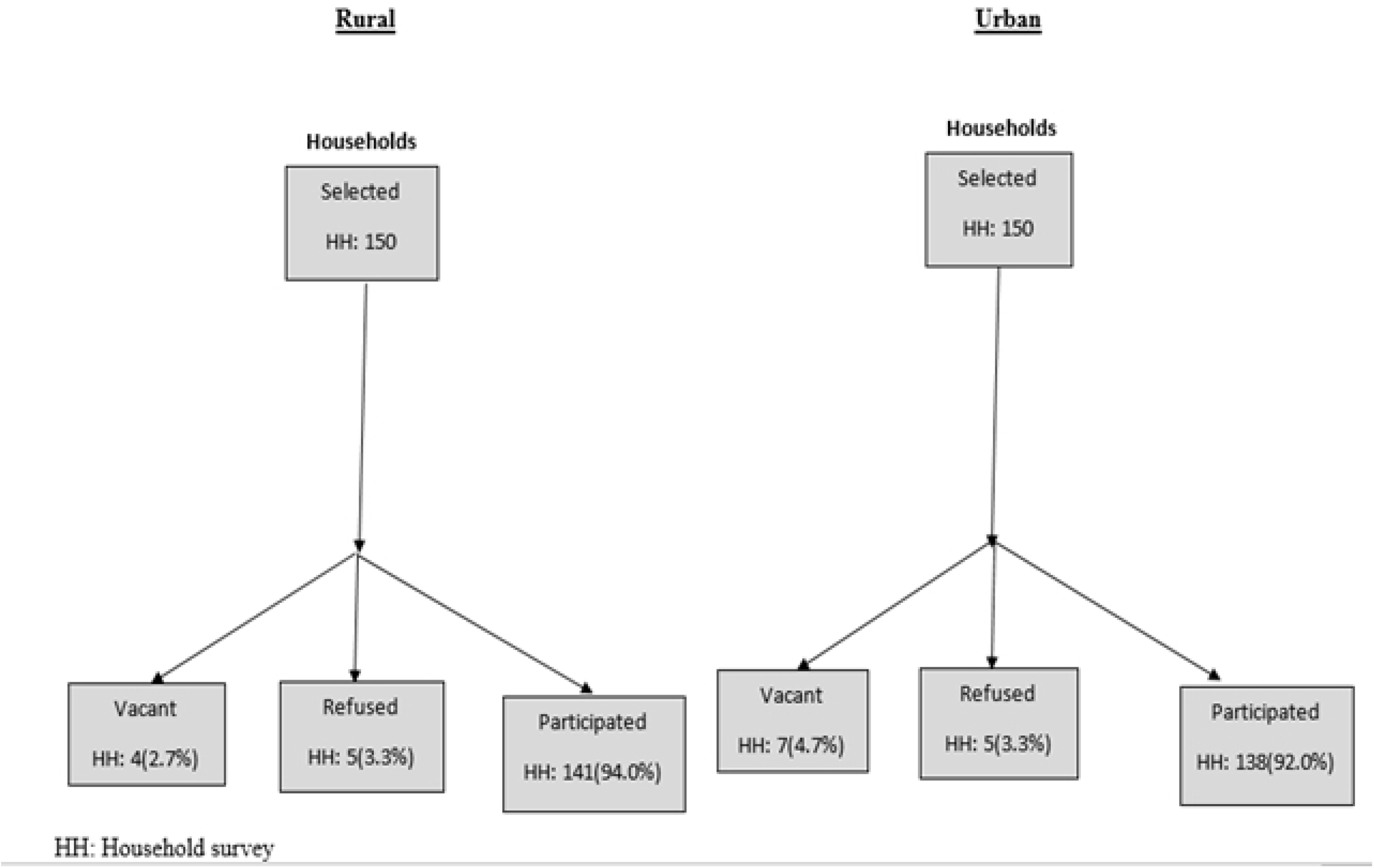
Study profile for households and participant’s selection.

The questionnaire captured demographic information, which included age, sex, occupation, level of education, marital status and religion, as well as socio-economic status, which includes house type, household size, and toilet type. Environmental and behavioral characteristics obtained using the questionnaire included presence of ceilings in houses, if windows are screened with nets, whether participants often stay out late into the night, presence of stagnant water around residences, crop cultivation around residences, presence of bushes around residences, LLINs usage, use of insecticide sprays, and living with animals in homes. People who had taken anti-malaria treatment less than two weeks before the survey were excluded from the study. A systematic sampling technique was used to select households for data collection and in recruiting participants in both rural and urban settlements of the Health District, a household was skipped after visiting the nearby house, until a total of 250 samples was achieved for each setting. The axillary body temperature was measured using an infrared thermometer (Manufactured by Medifriend, RoHS model, England-United Kingdom) and fever was defined as temperature ≥ 37.5 °C [36]. A finger prick was done using a sterile disposable lancet, to obtain a blood sample for laboratory analysis.

### Sample Size

The sample size was calculated using the 45.47% prevalence of malaria parasites in a study as reported by Lehman *et al*., 2018 in Douala. Sample size was determined using the formula n = Z^2^pq/d^2^ [29] where n = the sample size required, z = 1.96: which is the standard normal deviate for a 95% confidence interval, CI), p = 0.45 proportion of malaria prevalence, q = 1–p: proportion of malaria negative and d = acceptable error willing to be committed. The minimum sample size was estimated at n = 380. The sample size was then increased by 24% to a minimum of 500 participants to account for anticipated non-respondents, incomplete data entry and loss of samples due to blood clotting. A systematic sampling technique whereby a household was skipped after visiting the previous one was used to select households and 250 participants in rural areas and 250 participants in urban settlements of the Health District.

### Laboratory Analysis

Diagnosis of malaria was done by microscopy. Thick blood films were made from participant’s blood samples, air-dried and transported to Kaewo integrated health centre laboratory where they were stained with 5 % Giemsa for 25 minutes, rinsed, air dried and stored for onward transportation to the University of Buea life science laboratory for observation [11]. Each blood film was independently examined microscopically by two well-trained and well-experienced parasitologists, following standard procedure for the detection and identification of malaria parasites. The samples were observed under the light microscope at X100 objective (oil immersion). Slides were considered positive when schizonts, trophozoites and/or gametocytes of Plasmodium were observed on the blood film. A smear was declared negative, after observing 100 high power fields, and no malaria parasite morphology was seen. Positive slides were quantified by counting the number of parasites against 200 white blood cells and the parasites/μl blood calculated by assuming a leucocyte count of, 8000 per microliter as described elsewhere [11].

### Ethical considerations

Participants were informed on the potential benefit and aim of the study before obtaining their consent. Parents or guardians gave assent for minors (0-18 years) by filling out and signing the assent form. Ethical approval was obtained from the Faculty of Health Science Institutional Review Board of the University of Buea reference number: 2019/979-05/UB/SG/IRB/FHS. Administrative authorization was obtained from the Far North Regional Delegation of Public Health reference number: 374/AR/19/MINSANTE/SG/DRSP/EN/YT/MRA. Subsequent administrative authorizations were sought from village heads (Lawanats), quarter heads and community leaders of concerned localities. Written consent was obtained from each studied participant. For most of the participants who were unable to read or write French or English, the information was read and explained to them in Fulfulde which they best understood, and consent approval was indicated by thumb printing the consent form. Participants were given full right to participate or refuse participation in the study.

### Mosquito collection

We recruited 2 research assistants and one experienced Entomologist from the University of Buea. All the research assistants completed a two-day training course on data collection tools. Mosquitoes larvae and pupae were collected between August 2019 and October 2019 in Kaewo (rural area) and Dougoi (urban area), which were selected randomly (Fig 1). In each locality where there was stagnant water, breeding sites were identified and larvae were collected and reared locally by storing the stagnant water in buckets covered with a net, until adults emerged. The adults were fed with 10% glucose solution for 2-4 days before bioassay was conducted. Morphological identification was done using the identification criteria by [12] and *Anopheles* species were found to be dominant (>90%). Entomological inoculation rate (EIR) in these areas is estimated to range between 2.4–24.0 infective bites/person/month during the rainy season [37], and *Anopheles arabiensis* as the main vector species identified.

### Insecticide susceptibility bioassay

Mosquito’s resistance to LLINs in both rural and urban settlements of the health district were investigated using the CDC (Centers for Disease Control and Prevention) Bottle Bioassay test technique. The assay determines if the active chemical substance (insecticide used in LLINs-Pyrethroid) is able to kill mosquitoes from a specific location (rural and urban area of Maroua III health district) at a given time (30 minutes).

#### Insecticide sensitivity assay

The CDC Bottle Bioassay test kit consisting of 250ml Wheaton bottles, micropipettes, mouth aspirator, timer, titration flasks, and necessary insecticides were provided by CDC, 1600 Clifton Road. NE, Atlanta, GA, USA. The CDC bottle bioassay is an essential tool for detecting resistance to insecticides, during which five 250-ml Wheaton bottles with screw lids were washed with warm soapy water, rinsed thoroughly with water at least three times and air dried. After drying, the bottles and caps were marked with permanent stickers, with one of the bottles marked as control and the rest as test bottles (as shown in Fig 3 a and b). Using a pipette, 1ml of acetone was added into the control bottle. Using another pipette, 1 ml of the freshly prepared Deltamethrin stock insecticide solution (12.5 μg/mL in acetone solution) was added into the four test bottles. The bottles were caped and swirled until the interior of the bottles were completely coated. The bottles were then uncapped and allowed for 4 hours to completely dry. The bottles were left on their sides and protected from light before the introduction of mosquitoes for the experiment.

**Figure 3:**
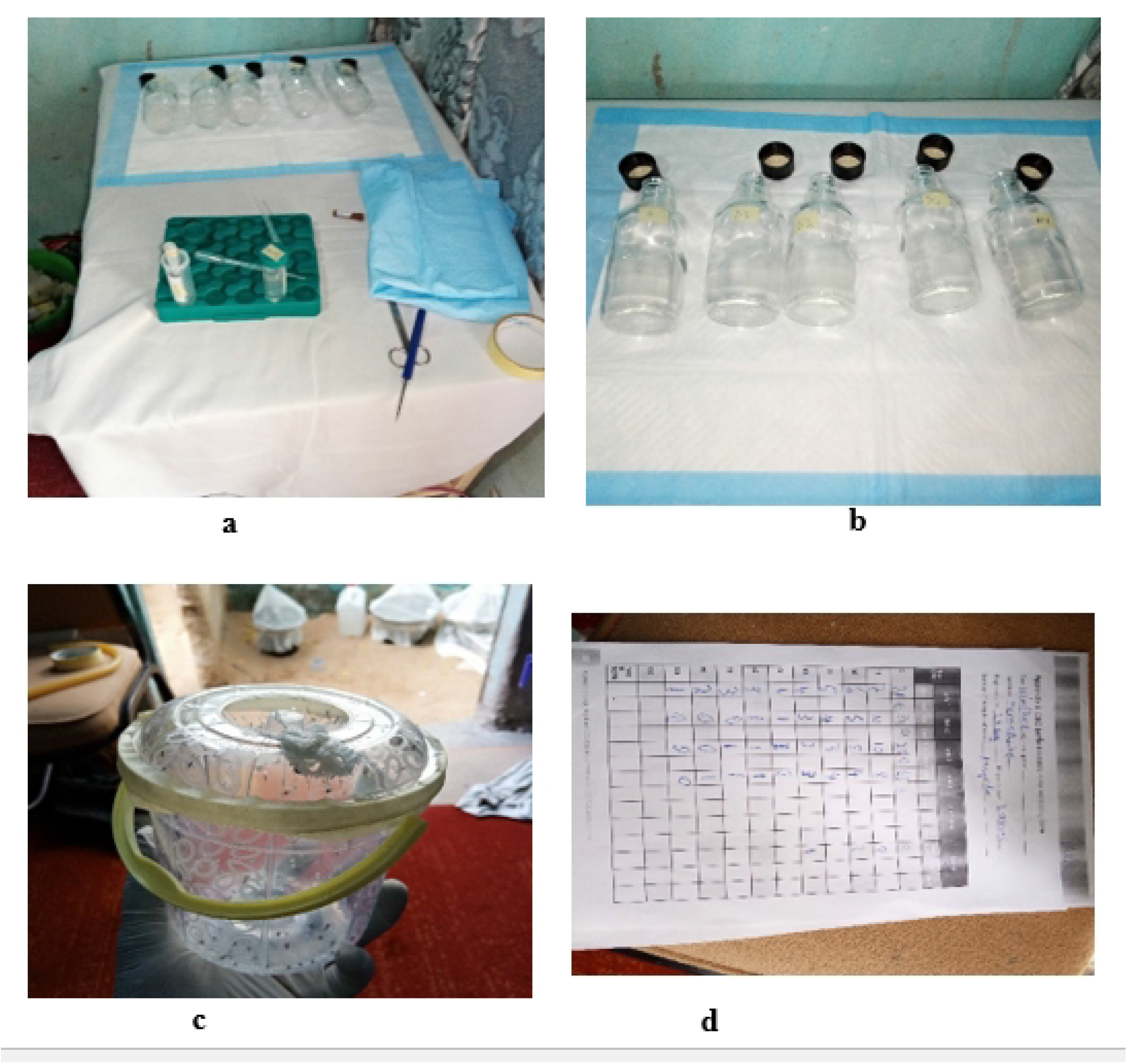
Pictorial presentation of CDC bioassay procedure by the research team. *End points* **a:** Tube of Deltamethrin, acetone and Wheaton bottles, **b:** Wheaton bottles impregnated with insecticide, **c:** mosquitoes within transparent bucket, **d:** record forms

Mosquitoes that were collected from the different health areas in the Health District (Fig 1) were used. The mosquitoes were first grown and fed with sugar solution for 3 days before the experiment (Fig 3 c). Using a mouth filter aspirator, between 10-40 mosquitoes (total of 96 mosquitoes) were gently blown into each bottle (control and test bottles). After filling the 5 bottles with the mosquitoes, the timer started and at Time 0, the number of dead and/or live mosquitoes were counted and recorded in an appropriate recording form (Fig 3 d). Dead and/or live mosquitoes were counted and recorded every 15 minutes for up to 2 hours, which marked the end of the experiment. Mosquitoes were considered dead when they could no longer stand to fly. Graphing of the total percentage mortality (Y axis) against time (X axis) was done on a linear scale. During the investigation, the diagnostic time (30 minutes) was the most critical value because it represents the threshold between susceptibility and resistance. The procedure was repeated using 21.5 μg/mL of permethrin, dissolved in acetone solution.

Reference diagnostic doses and diagnostic time for the insecticides used were 12.5ug/ml in 30 minutes for Deltamethrin and 21.5 ug/ml in 30 minutes for permethrin to achieve 100% mortality, against which results were compared. Resistance was assumed to be present if a portion of the test population survived the diagnostic dose at the diagnostic time (30 minutes). If test mosquitoes survive beyond this threshold, these survivors represent a proportion of the population that is resistant to the insecticide. All mosquitoes that died before the diagnostic time, after exposure to the insecticide-coated bottles, were considered as susceptible.

### Statistical Analysis

Data collected was cleaned up and analysed using the IBM-Statistical Package for Social Sciences (IBM-SPSS) version 25. Categorical variables reported as frequencies and percentages were used to evaluate the descriptive statistics. A p-value < 0.05 was considered significant. The prevalence of malaria in both rural and urban areas of the Health District was computed using the formula;

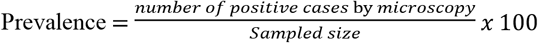

Logistic regression analysis was used to identify risk factors associated with malaria by comparing demographic factors, socioeconomic status, behavioral factors and environmental factors as independent variables against the presence or absence of malaria infection as dependent variables. Bivariate logistic regression was used initially to identify significant risk factors at p-value <0.05, which was confirmed using multivariate regression analysis. Crude and adjusted Odds Ratios (OR) as well as their 95% Confidence Intervals (CI) were computed for comparative analysis of rural and urban settlements in the Health District. The Mann-Withney U test was used to compute geometry mean parasite density (GMPD) as the test variable against gender for the studied participants. The Kruskal–Wallis H test was used to compute GMPD as the test variable against other demographic variables like age, occupation and marital status. QtiPlot 0.9.8.3 was used to compute a graph of resistance analysis, in which percentage mortality in mosquitoes was plotted against time, to determine the mean mortality rate and the result was compared with the WHO standard for resistance monitoring which states that a mortality of 98 to 100% at the recommended diagnostic time indicate susceptibility.

## RESULTS

### Prevalence of malaria in Maroua III Health District

Of the 500 blood samples examined using microscopy, the prevalence of malaria in Maroua III health district was 52.2%. The prevalence was significantly higher (p=0.016) in rural areas (57.6%) as compared to urban areas (46.8%), Fig 4. Asymptomatic malaria parasitemia (participants with malaria presenting with no signs and symptoms of malaria) was (49.6%) in rural areas. This was more than quadruple symptomatic malaria (8%). Likewise, this was similar in urban areas, where asymptomatic malaria parasitemia (37.2%) was also higher than symptomatic malaria infection (9.6%). The prevalence of asymptomatic malaria parasitemia was significantly higher (p = 0.029) in rural areas than in urban settings. In the entire district, the prevalence of asymptomatic malaria parasitemia was 43.4% as compared to symptomatic malaria which was 8.8 % and 3.2% (16) of the participants who were not infected with malaria had fever with temperatures greater than 37.6^0^C

**Fig 4:**
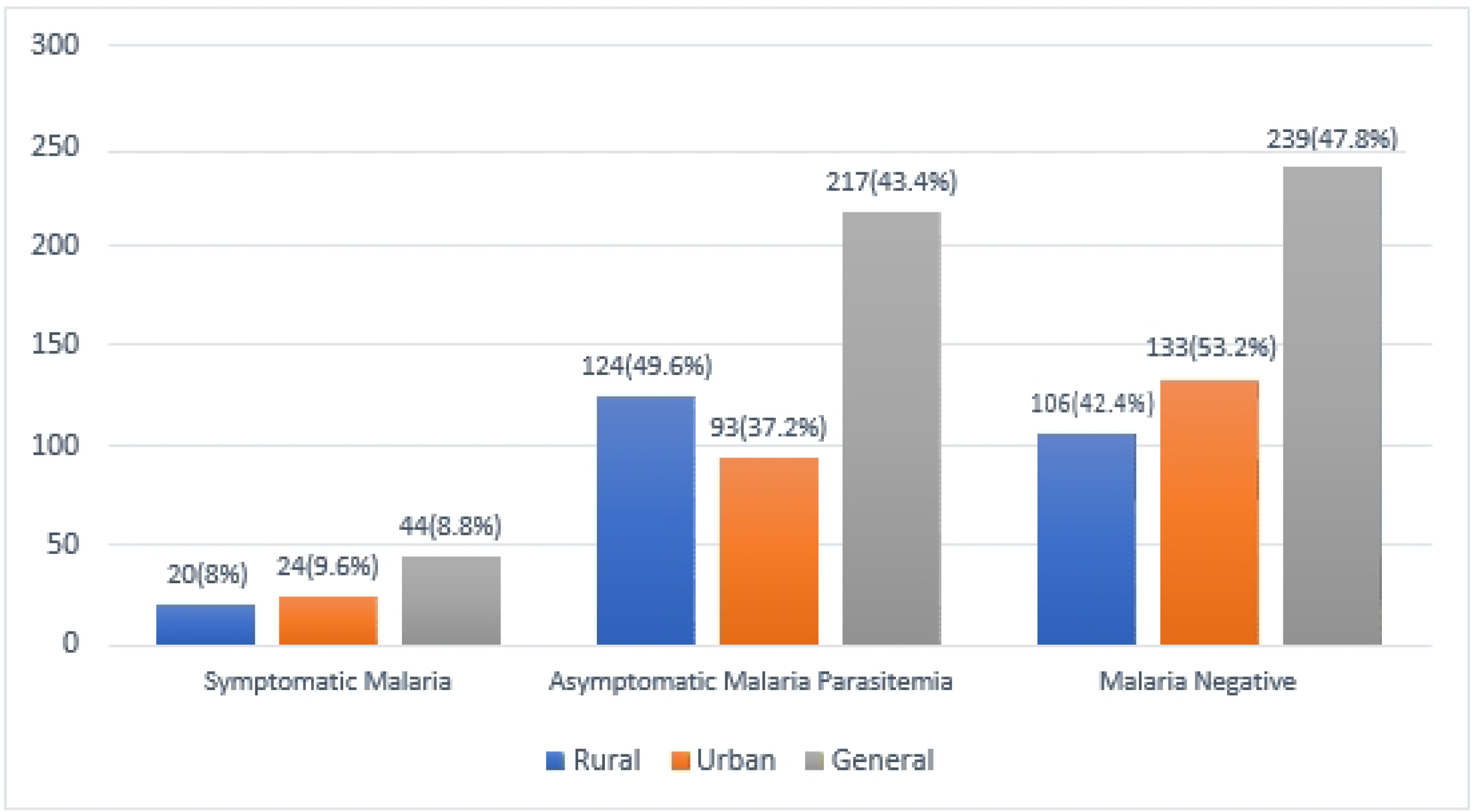
Prevalence of malaria in Maroua III Health District. Definitions and endpoints ➢ Asymptomatic malaria was defined as the presence of Plasmodium with an axillary temperature of < 37.5 °C. ➢ Symptomatic malaria parasitemia was defined as the presence of Plasmodium, with an axillary temperature of ≥ 37.5 °C, joint pains, vomiting, headache, diarrhea, chills

### Association of malaria infection with demographic factors

From the Chi-square test, the prevalence of malaria was significantly associated with age (p = 0.025), occupation (0.041) and marital status (0.003) in rural areas of the district as shown in Table 1. Meanwhile, malaria prevalence in urban areas was significantly associated with age (p < 0.001), educational level (p = 0.002), occupation (p < 0.001), marital status (p <0.001) and religion (p = 0.005). Malaria infection was more common among children 64(25.6%) than any other age group in rural communities, as well as among children 45(18.0%) in urban settings. Equally, malaria infection was highest among pupils, 72(64.9%) as compared to any other occupational group in the rural communities. Likewise, in urban areas, 55(56.7%). Table 1 also shows that about two thirds of participants in both rural and urban areas had primary school education only. In the urban area, many more participants had a university education compared to rural communities.

**Table 1:**
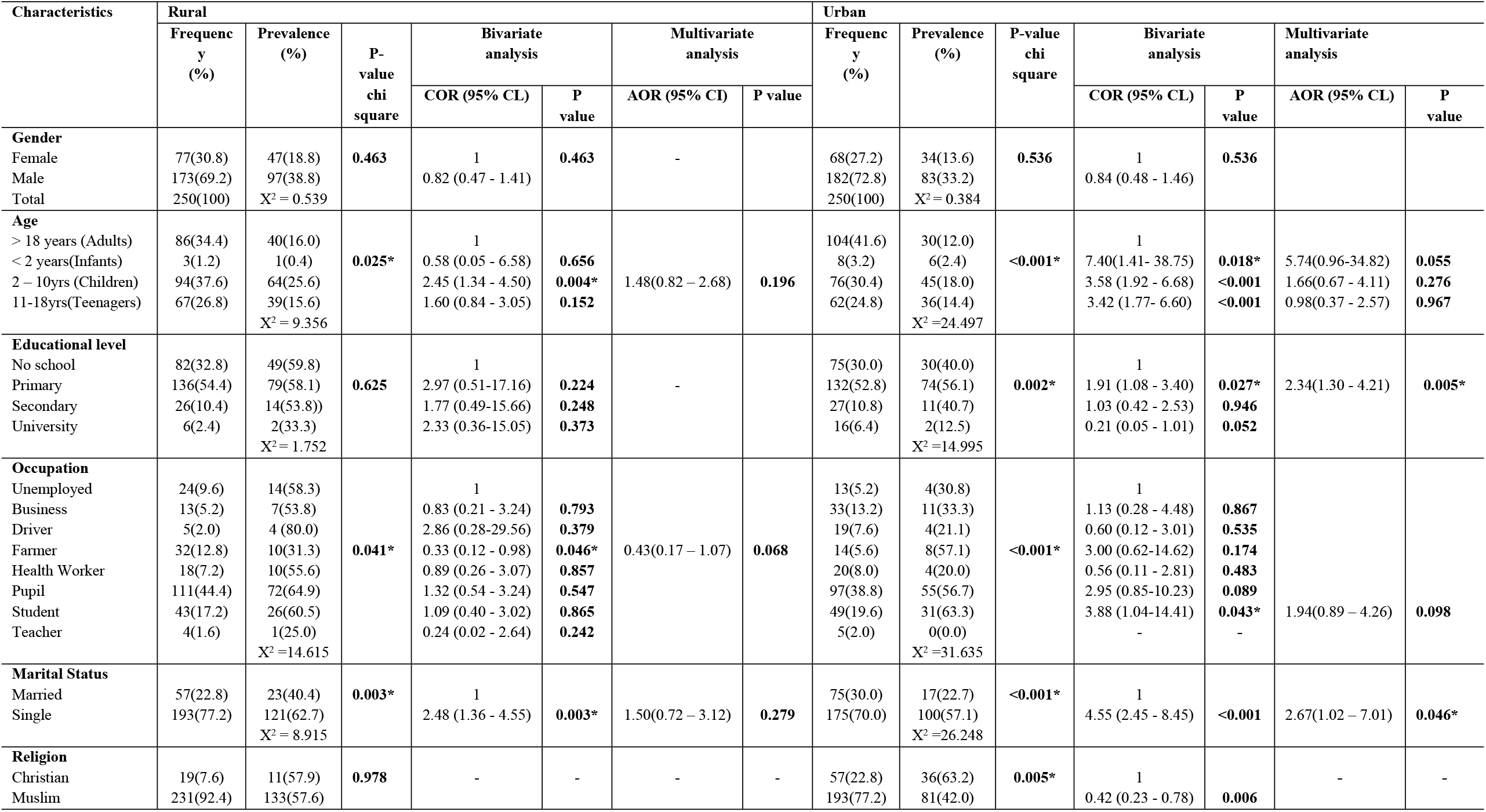
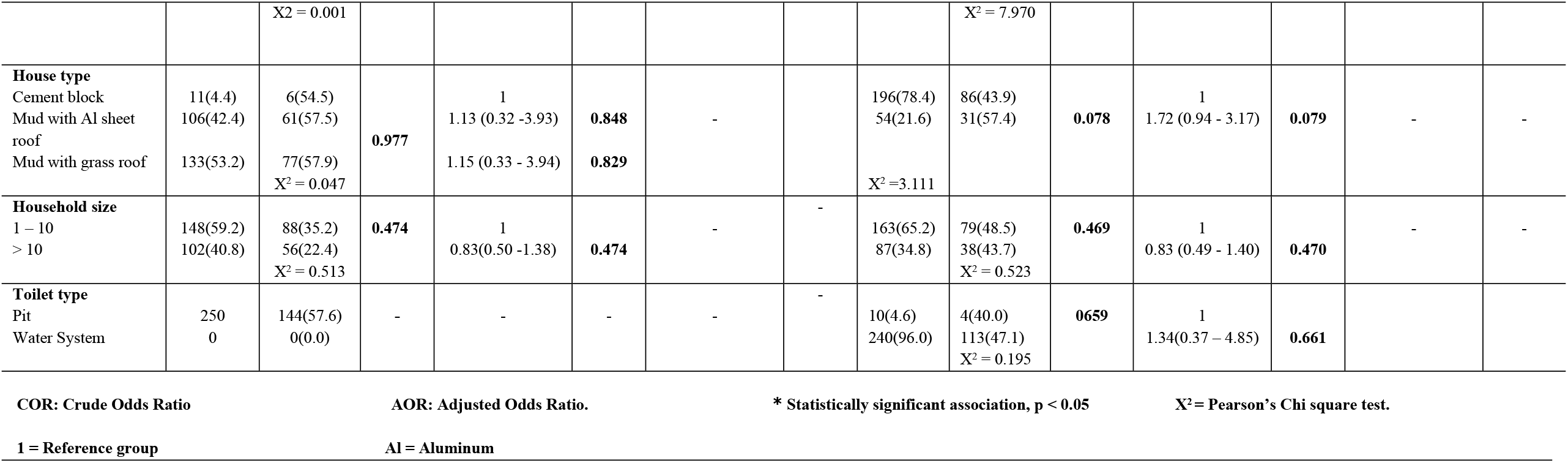
Malaria Infection in Rural and Urban areas of Maroua III Health District in relation to demographic and socioeconomic factors.

Logistic regression analysis was carried out between malaria prevalence as dependent variable and sociodemographic factors as independent variables, for all the variables which were significant by bivariate analysis. In the bivariate analysis age group 2-10years (cOR = 2.45, 95% CI =1.34 -4.50, p = .004), being a farmer (cOR = 0.33, 95% CI = 0.12 – 0.98, p = .046), and being single (cOR = 2.48, 95% CI =1.36 -4.55 p = 0.003) were significantly associated with malaria infection among inhabitants of Maroua III district in rural areas. Meanwhile, in urban settlements for the bivariate analysis, infants, children, teenagers (cOR = 7.40, 95% CI =1.41 – 38.75, p = .018; cOR = 3.58, 95% CI =1.92 – 6.68 p < 0.001; cOR = 3.42, 95% CI =1.77 - 6.60, p <0.001), being at primary (cOR = 1.91, 95% CI =1.08 – 3.40, p = .027) and being a student in secondary school (cOR = 3.88, 95% CI =1.04 – 14.41, p = .043) were significantly associated with a higher prevalence of Malaria infection. After performing the multivariate logistic regression analysis using the bivariate model of significant variables p<0.05, primary school children (p = 0.005), and being single (0.046) were found to be the significant risk factors of malaria among the people in urban settlements of Maroua III health district. No demographic factor in rural areas was significant in the multivariate model. On the whole, for the entire health district, multivariate analysis showed that the age group 2-10years had 2.75 odds of contracting malaria over adults 18 years and above. Likewise, teenagers 11-18years had 2.26 odds of contracting malaria over adults 18 years and above (Table 4)

### Environmental and behavioral characteristics of the study population in relation to malaria infection

Malaria risk factors in rural areas of Maroua III health district were quantified using logistic regression to calculate bivariate and multivariate odds ratios with their respective confidence intervals as in Table 2. According to bivariate analysis, significant odds ratio for malaria in rural areas within the population of Maroua III health district were; participants sleeping on beds with LLINs older than 3 years (cOR: 2.17, 95% CI: 1.18–4.00, p = 0.013) over households with LLIINs less than 3 years old and households which source of water is from opened/unprotected wells (cOR:1.55, 95% CI: 1.05–6.22, p 0.039) over those households which source of water are from well-constructed wells “Forage”.

**Table 2:**
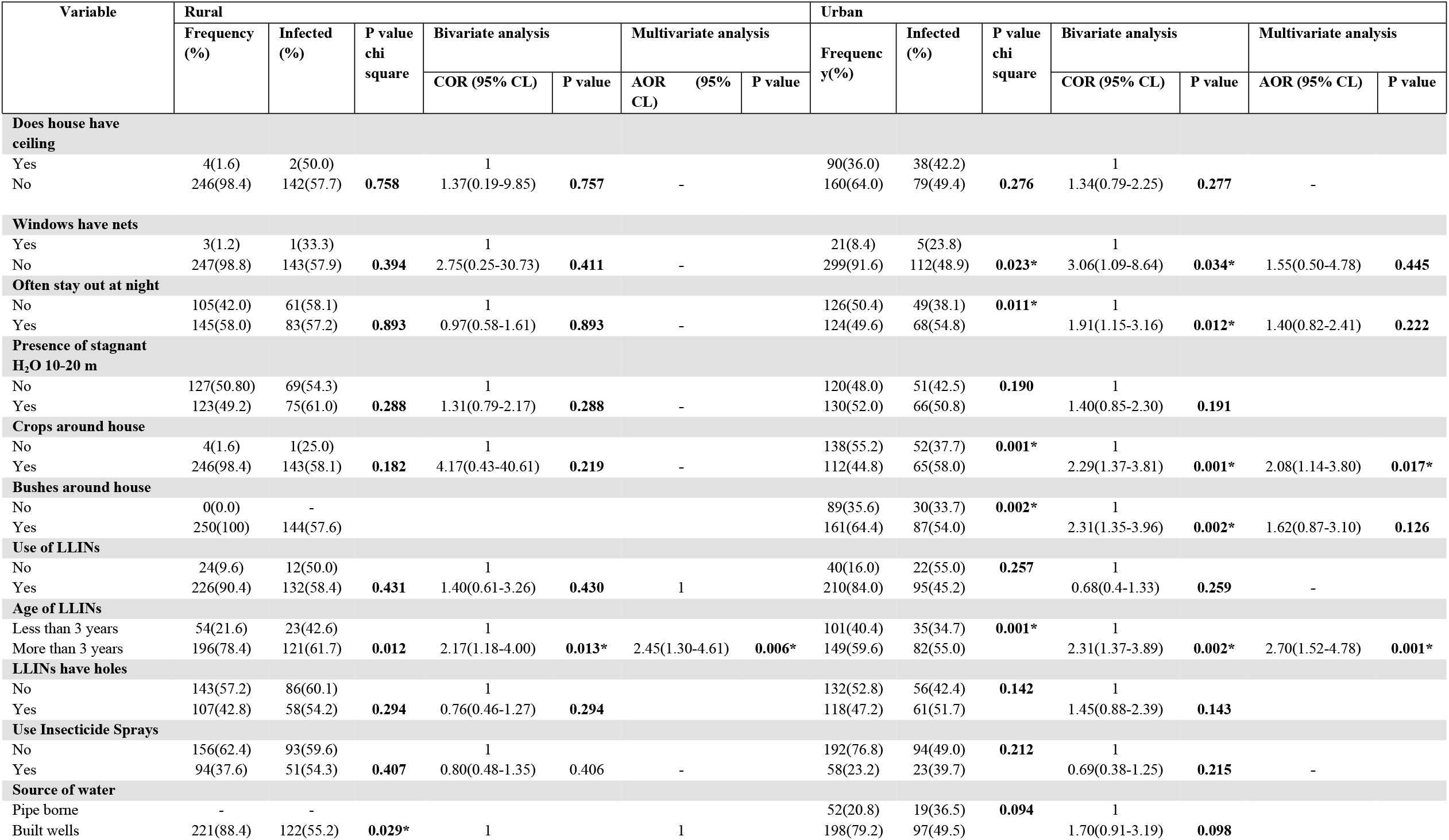
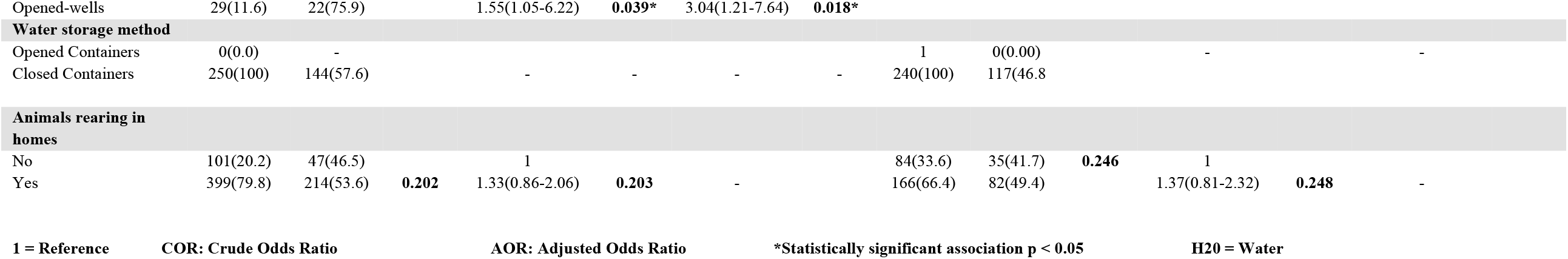
Environmental and Behavioral characteristics of the study population in relation to malaria infection in rural and urban areas of Maroua III HD.

**Table 3:**
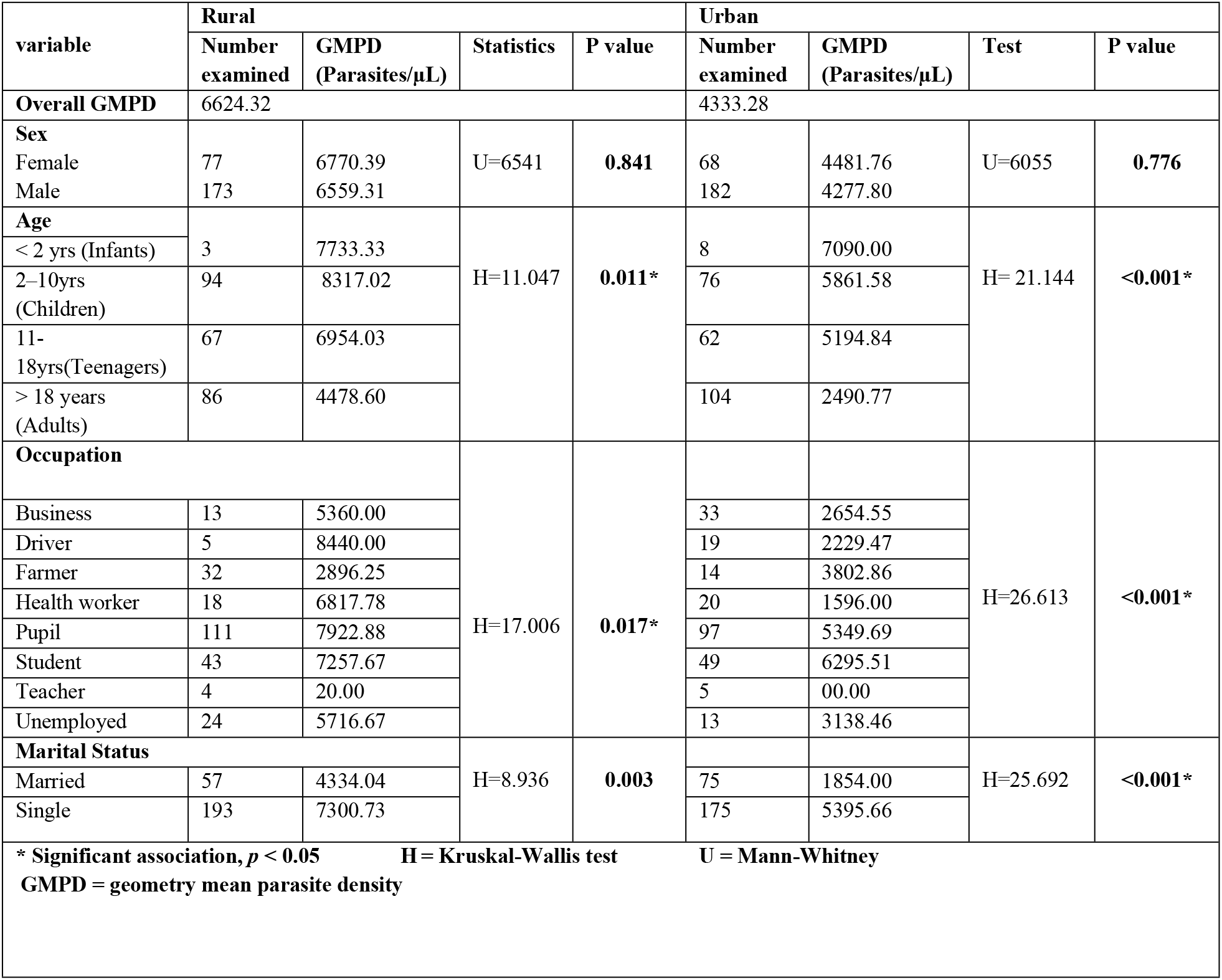
Association of Geometric mean parasite density (GMPD) with demographic factors.

According to the multivariate analysis, the following risk factors were statistically significant in the rural area: Participants who were using mosquito bed nets (LLINs) which were obtained more than three years ago had 1.45 increased odds of contracting malaria than those who were using mosquito bed nets obtained less than three years ago. Participants whose sources of water are opened and unconstructed wells had 3.04 increased odds of contracting malaria.

Likewise, malaria risk factors in the urban areas of Maroua III health district were identified in a similar manner as done for the rural area (Table 2). According to the bivariate analysis, the following odds ratios were reported: Those households that do not have mosquito nets on their windows (cOR: 3.06, 95% CI: 1.09–8.64, p = 0.034) over households who had mosquito nets on their windows. Participants who stay out late into the night (cOR: 1.91, 95% CI: 1.15–3.16, p = 0.012) over those who do not stay out late into the night. Those households surrounded by farmland and bushes (cOR: 2.29, 95% CI: 1.37–3.81, p = 0.001; cOR: 2.31, 95% CI: 1.35–3.96, p = 0.002) over households not surrounded by farmland and bushes. Those households who obtained their bed nets more than three years ago (cOR: 2.31, 95% CI: 1.37–3.89, p = 0.002) over households who had obtained bed nets less than three years ago.

All the significant factors identified in the bivariate analysis were retained and included in the multivariate model. According to the multivariate logistic regression analysis, the following risk factors were statistically significant in the urban areas of the district. Participants who live in households surrounded by cultivated farm land had 2.08 odds of contracting malaria over those who live in households not close to farmland. Households who use LLINs older than 3 years had 2.70 odds of contracting malaria over households that used LLINs obtained less than 3 years ago.

On the whole, for the entire district, farmland around residences and using LLINs older than 3 years were significant environmental factors associated with malaria as shown in Table 5.

**Table 4:**
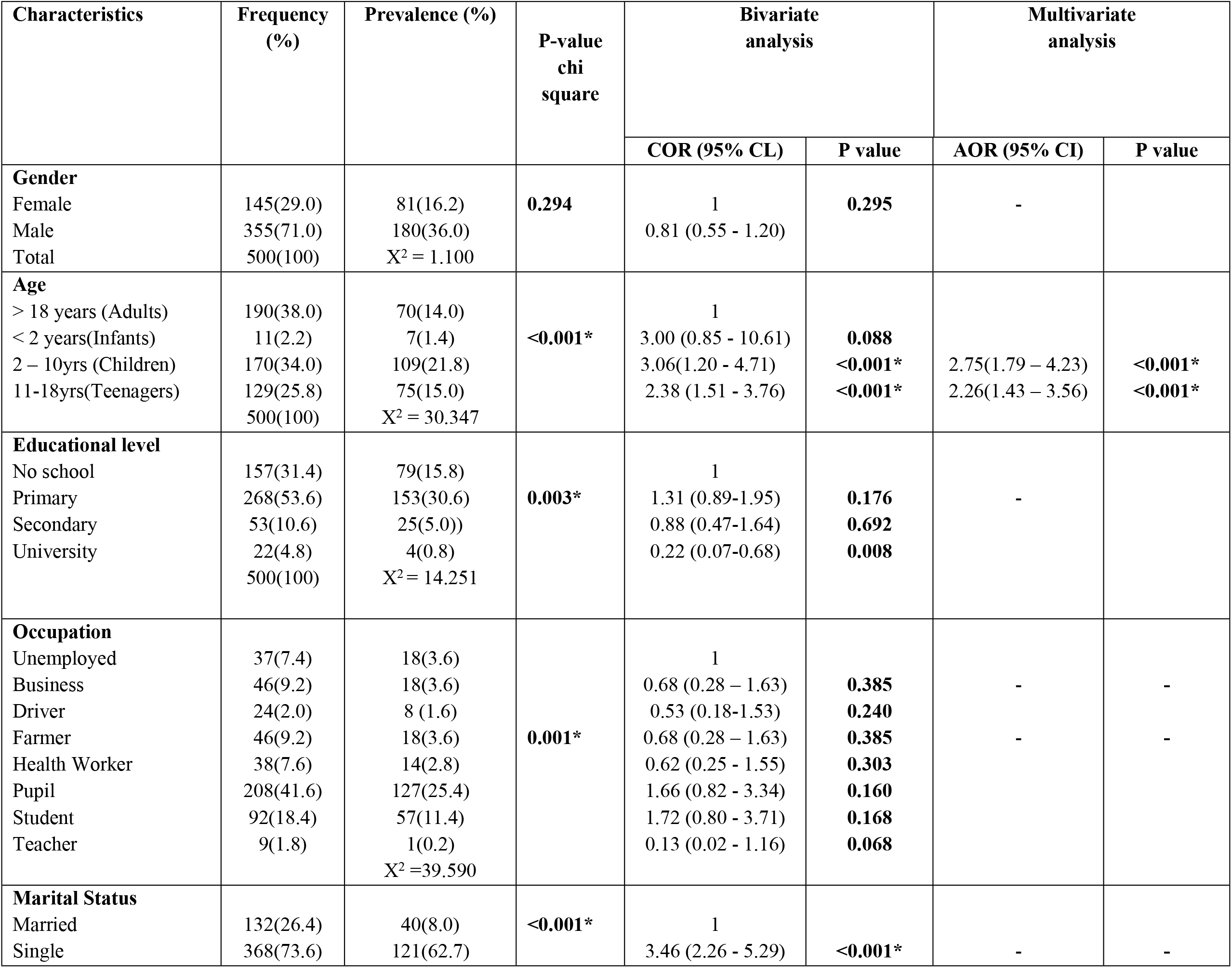
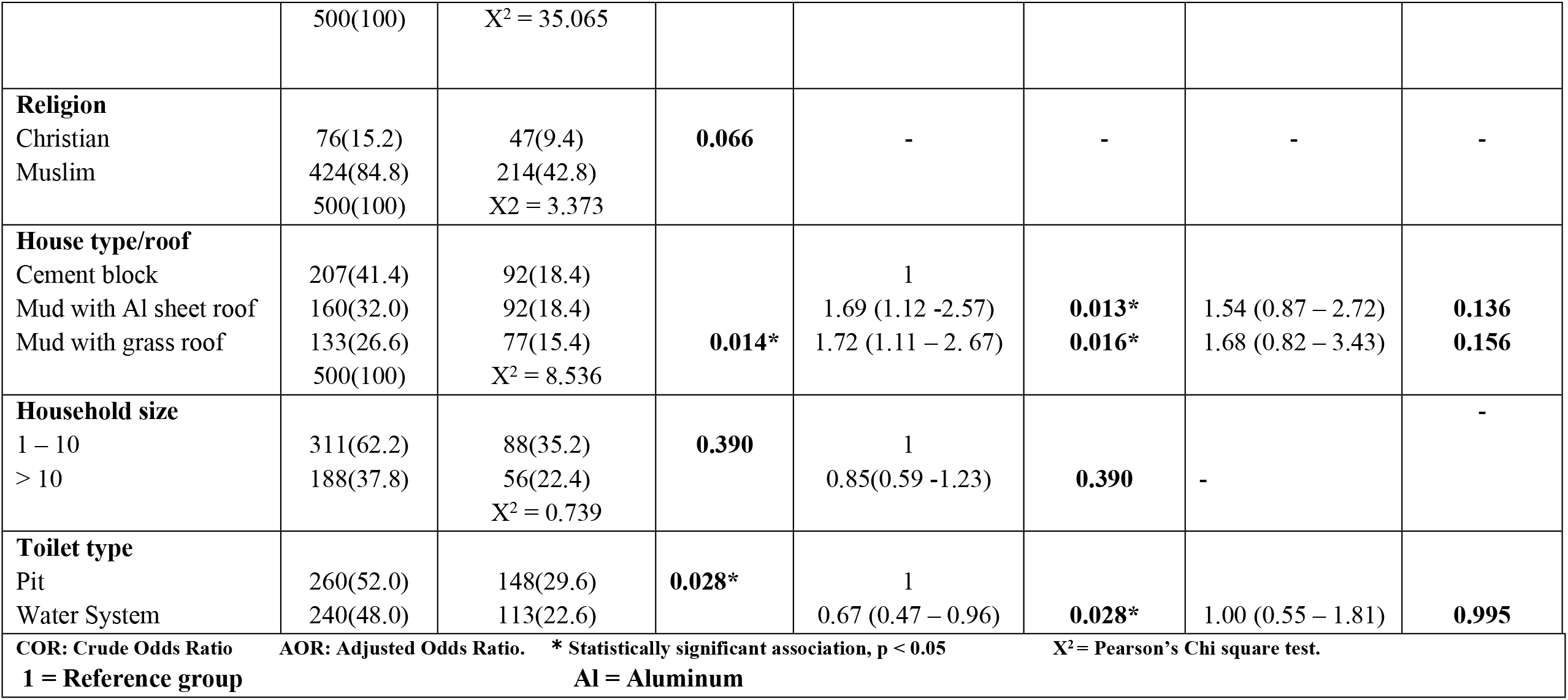
Malaria Infection in Maroua III Health District in relation to demographic and socioeconomic factors.

**Table 5:**
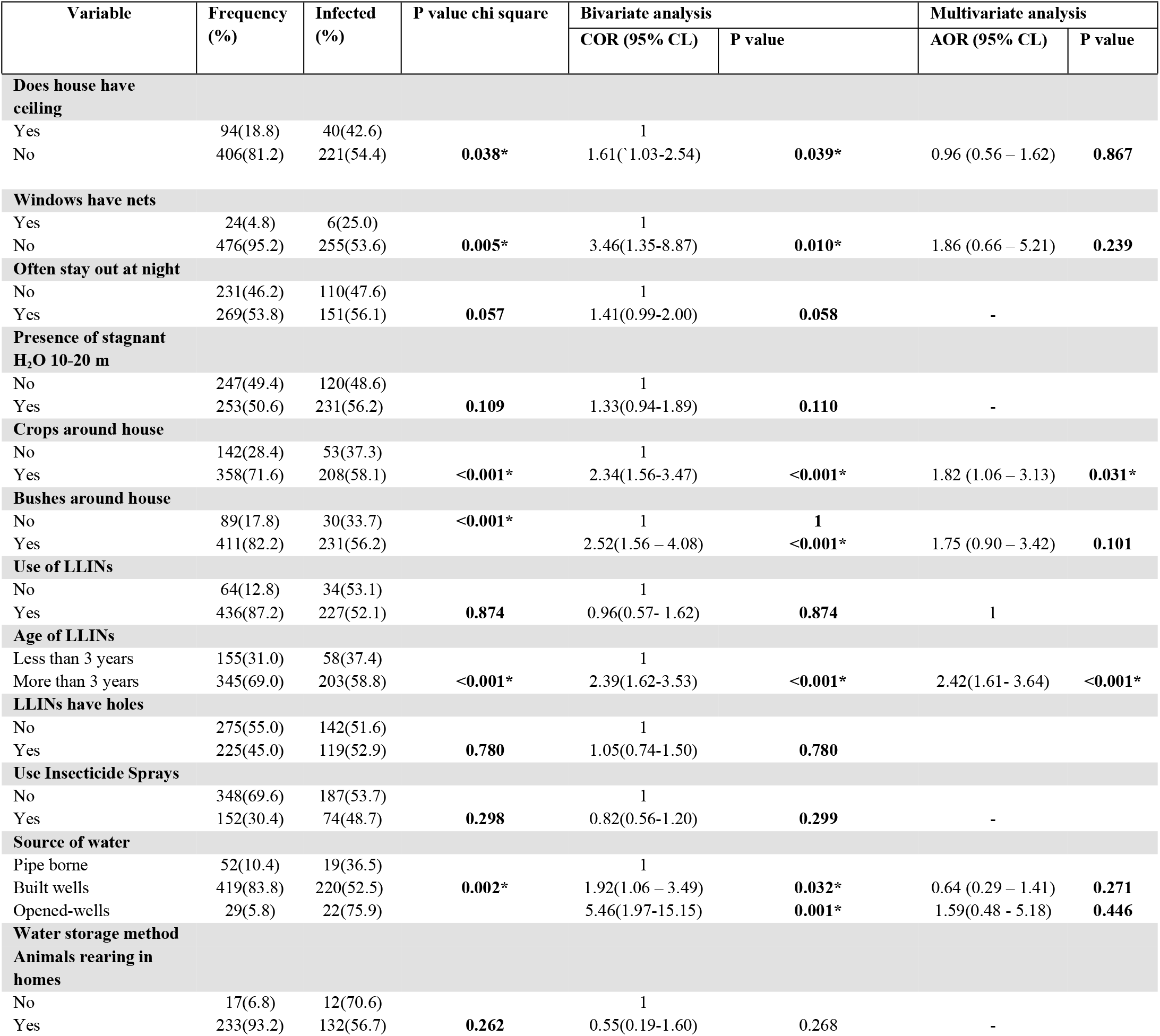
Environmental and Behavioral characteristics of the study population in relation to malaria infection in Maroua III HD as a whole.

| Variable | Frequency (%) | Infected (%) | P value chi square | Bivariate analysis |  | Multivariate analysis |  |
| --- | --- | --- | --- | --- | --- | --- | --- |
|  |  |  |  | COR (95% CL) | P value | AOR (95% CL) | P value |
| Does house have ceiling |  |  |  |  |  |  |  |
| Yes | 94(18.8) | 40(42.6) |  | 1 |  |  |  |
| No | 406(81.2) | 221(54.4) | 0.038* | 1.61(1.03-2.54) | 0.039* | 0.96 (0.56 – 1.62) | 0.867 |
| Windows have nets |  |  |  |  |  |  |  |
| Yes | 24(4.8) | 6(25.0) |  | 1 |  |  |  |
| No | 476(95.2) | 255(53.6) | 0.005* | 3.46(1.35-8.87) | 0.010* | 1.86 (0.66 – 5.21) | 0.239 |
| Often stay out at night |  |  |  |  |  |  |  |
| No | 231(46.2) | 110(47.6) |  | 1 |  |  |  |
| Yes | 269(53.8) | 151(56.1) | 0.057 | 1.41(0.99-2.00) | 0.058 | - |  |
| Presence of stagnant H<sub>2</sub>O 10-20 m |  |  |  |  |  |  |  |
| No | 247(49.4) | 120(48.6) |  | 1 |  |  |  |
| Yes | 253(50.6) | 231(56.2) | 0.109 | 1.33(0.94-1.89) | 0.110 | - |  |
| Crops around house |  |  |  |  |  |  |  |
| No | 142(28.4) | 53(37.3) |  | 1 |  |  |  |
| Yes | 358(71.6) | 208(58.1) | <0.001* | 2.34(1.56-3.47) | <0.001* | 1.82 (1.06 – 3.13) | 0.031* |
| Bushes around house |  |  |  |  |  |  |  |
| No | 89(17.8) | 30(33.7) | <0.001* | 1 | 1 |  |  |
| Yes | 411(82.2) | 231(56.2) |  | 2.52(1.56 – 4.08) | <0.001* | 1.75 (0.90 – 3.42) | 0.101 |
| Use of LLINs |  |  |  |  |  |  |  |
| No | 64(12.8) | 34(53.1) |  | 1 |  |  |  |
| Yes | 436(87.2) | 227(52.1) | 0.874 | 0.96(0.57- 1.62) | 0.874 | 1 |  |
| Age of LLINs |  |  |  |  |  |  |  |
| Less than 3 years | 155(31.0) | 58(37.4) |  | 1 |  |  |  |
| More than 3 years | 345(69.0) | 203(58.8) | <0.001* | 2.39(1.62-3.53) | <0.001* | 2.42(1.61- 3.64) | <0.001* |
| LLINs have holes |  |  |  |  |  |  |  |
| No | 275(55.0) | 142(51.6) |  | 1 |  |  |  |
| Yes | 225(45.0) | 119(52.9) | 0.780 | 1.05(0.74-1.50) | 0.780 |  |  |
| Use Insecticide Sprays |  |  |  |  |  |  |  |
| No | 348(69.6) | 187(53.7) |  | 1 |  |  |  |
| Yes | 152(30.4) | 74(48.7) | 0.298 | 0.82(0.56-1.20) | 0.299 | - |  |
| Source of water |  |  |  |  |  |  |  |
| Pipe borne | 52(10.4) | 19(36.5) |  | 1 |  |  |  |
| Built wells | 419(83.8) | 220(52.5) | 0.002* | 1.92(1.06 – 3.49) | 0.032* | 0.64 (0.29 – 1.41) | 0.271 |
| Opened-wells | 29(5.8) | 22(75.9) |  | 5.46(1.97-15.15) | 0.001* | 1.59(0.48 - 5.18) | 0.446 |
| Water storage method |  |  |  |  |  |  |  |
| Animals rearing in homes |  |  |  |  |  |  |  |
| No | 17(6.8) | 12(70.6) |  | 1 |  |  |  |
| Yes | 233(93.2) | 132(56.7) | 0.262 | 0.55(0.19-1.60) | 0.268 | - |  |

### Parasite density of Malaria in rural and urban settlements of Maroua III HD

The geometry mean parasite density (GMPD) for rural settlement was 6624 parasites/μL of blood, which was higher than that of urban settlement (4333 parasites/μL of blood) and for the entire health district (5479 parasites/μL of blood) as shown in Table 3. The Kruskal-Wallis H test showed that the geometric mean parasite density was significantly different between age groups in both rural(p=0.011) and urban (p<0.001) settlements. Also, the geometric mean parasite density was significantly different (p = 0.017) amongst participants in rural settlements with pupils having the highest geometric mean parasite density (7923 parasites/μL of blood) compared to other occupations. Equally, the geometric mean parasite density was significantly different (p <0.001) amongst participants in urban settlement with students having the highest geometric mean parasite density (6296 parasites/μL of blood) compared to other occupations. In regard to marital status, the GMPD was significantly (*p* = 0.003) higher among singles (7301 parasites/μL of blood) than those married (4334 parasites/μL of blood), (H = 15.420/μL), in rural settlements. Likewise, this strength was the same for singles (5396 parasites/μL of blood) than those married (1854 parasites/μL of blood) in urban settlements. The geometric mean parasite density was found to decrease significantly (p<0.001) with increasing age of participants in both rural and urban settlements of the district, as shown in Fig 5.

**Fig 5.**
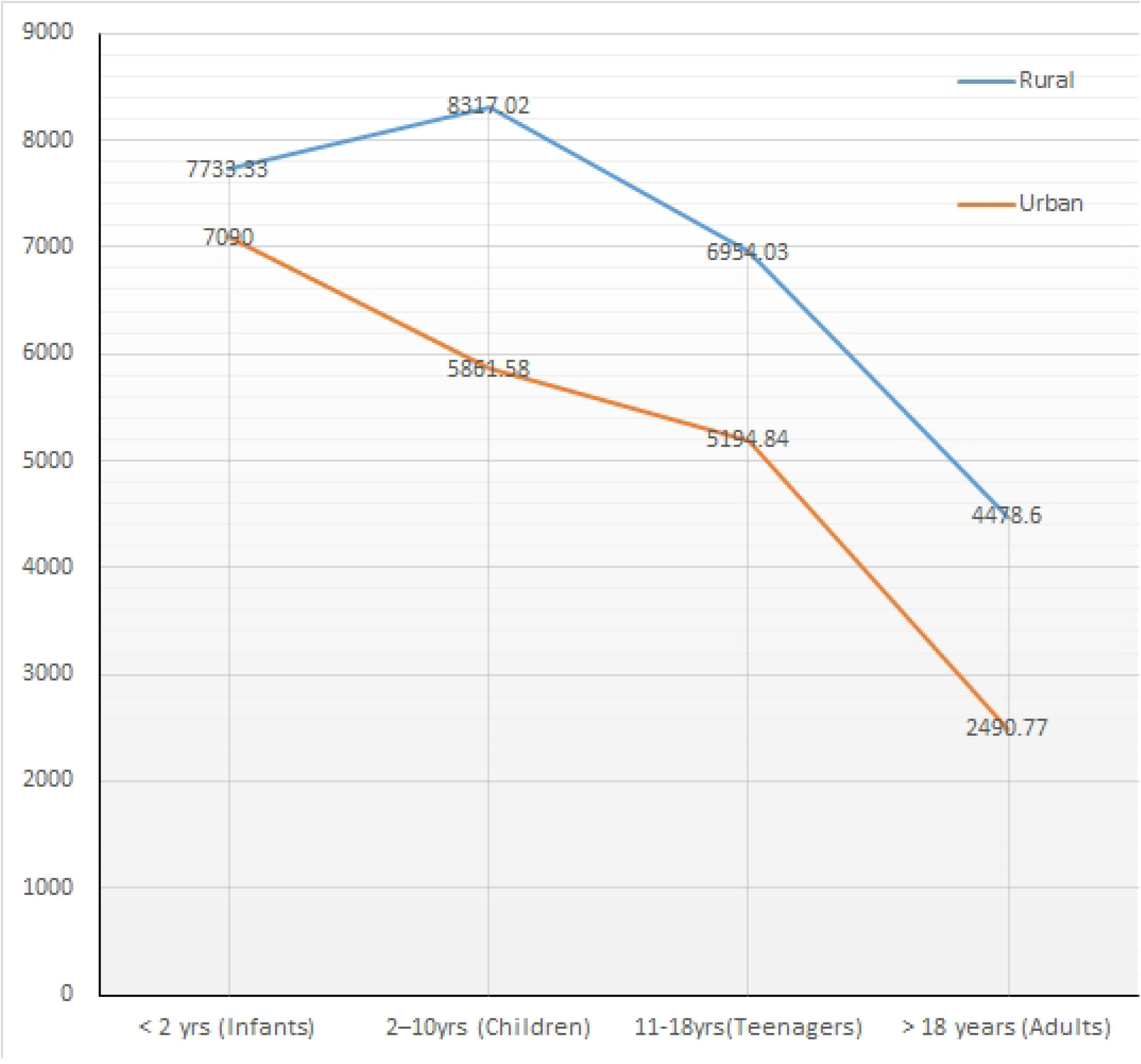
Parasite density strength with respect to age groups in rural and urban settlements of Maroua III HD.

### Effect of Deltamethrin on *Anopheles* mosquitoes obtained from Maroua III Health District

In Kaewo (rural area), 91.19% of the *Anopheles* species were susceptible to Deltamethrin insecticide and 8.81% were resistant, after the diagnostic time of 30 minutes as shown in Fig 6. Contrary to what obtains in Kaewo, a greater percentage of susceptible mosquitoes to Deltamethrin (95.83%) was observed in Dougoi (urban area) and 4.18% (6 mosquitoes out of 96) were resistant after the diagnostic time of 30 minutes. On a whole, 93.51% of mosquito susceptibility to Deltametrin was observed for the entire health district. The percentage of mosquitoes from Kaewo and Dougoi susceptible to Deltamethrin was within the WHO range of 80 to 97% mortality, which is interpreted as ‘possibility of resistance that needs more investigations to be confirmed’.

**Fig 6:**
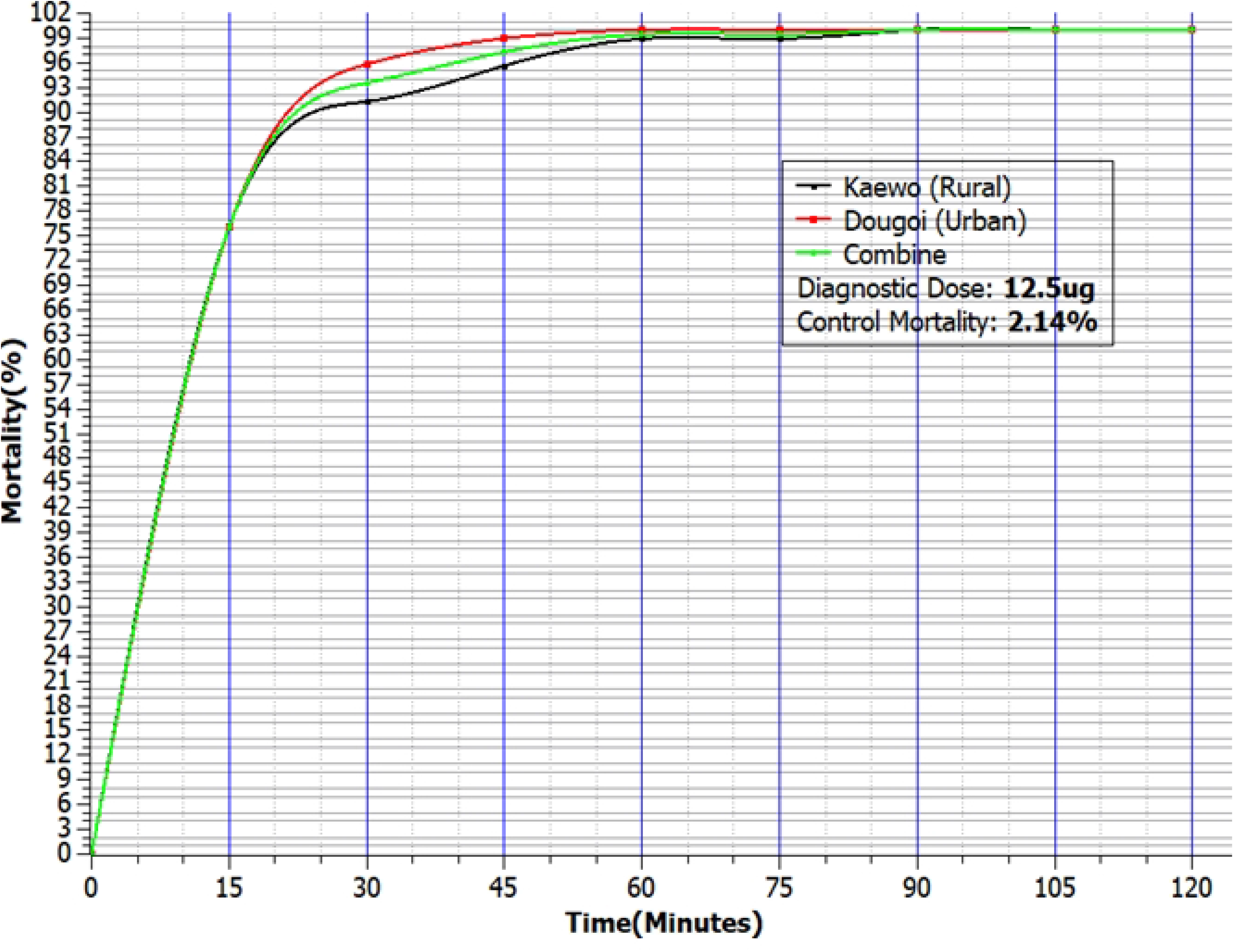
Mortality of *Anopheles* species mosquitoes observed after two hours of exposure to CDC bioassay bottles treated with Deltamethrine in Kaewo and Dougoi health areas of Maroua III HD.

### The Effect of Permethrin on mosquitoes obtained from Maroua III Health District

Mosquitoes obtained from Kaewo showed 85.24% susceptibility to permethrin and 14.76% resistance after the diagnostic time of 30 minutes, Fig 7. However, a lower susceptibility of mosquitoes (82.46%) was observed in Dougoi and 17.54% of the mosquitoes were resistant to permethrin at the diagnostic time of 30 minutes. On the whole, a percentage susceptibility of 83.85 was observed for the Maroua III health district. These percentages of susceptibility are within the WHO range of 80 to 97% mortality, which is interpreted as ‘possibility of resistance that needs to be confirmed’.

**Figure 7:**
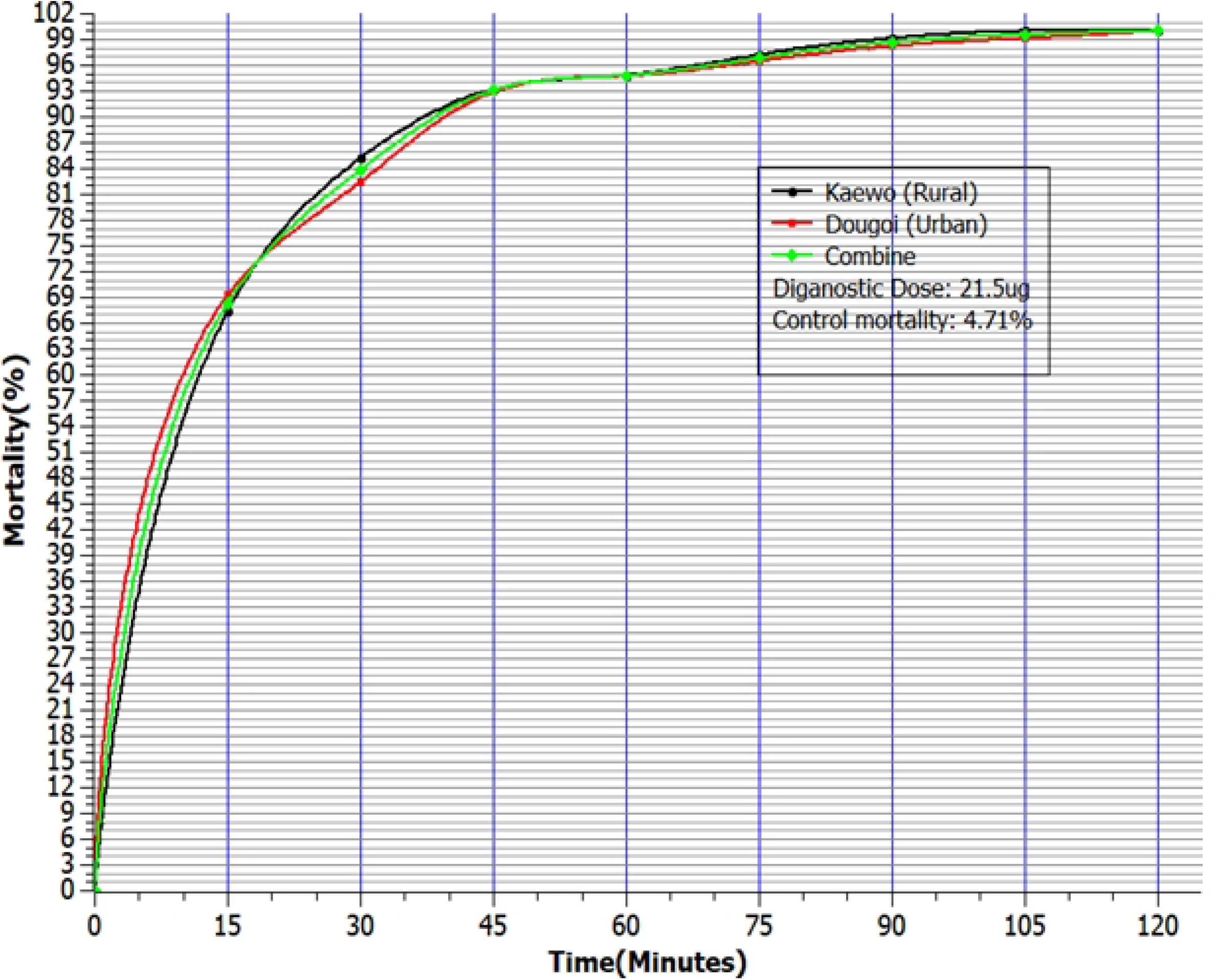
Mortality of *Anopheles* species mosquitoes observed after two hours of exposure to CDC bioassay bottles treated with Permethrin in Kaewo and Dougoi health areas of Maroua III HD.

## DISCUSSION

This investigation, which was carried out in the Sahel area, with seasonal malaria, revealed an overall malaria infection prevalence of 52.2 %, which was higher than that reported in Douala (45.47%), the economic capital of Cameroon [13], despite ongoing control measures. The prevalence of malaria was found to be significantly (p=0 .016) higher in rural areas (57.6%) than in urban settings (46.8%). This is in agreement with the observation that the level of malaria transmission in any area is generally higher in rural than in urban settings due to environmental factors [14, 15]. Generally, it is considered that suitable breeding sites are scarce in highly populated urban areas, and this leads to a reduction in the frequency and transmission dynamics of malaria. However, evidence of the adaptation of malaria vectors to the African urban environment has been reported in the past [16]. This work was carried out in the rainy season, which is a high transmission season with an increased number of breeding sites in these areas, leading to an increase in vector density with high inoculation rates and consequently, higher prevalence of malaria infection. Past studies have also reported seasonal variation in malaria prevalence, which was higher during the rainy season than in the dry season [17, 18].

It was revealed in this study that malaria infection was significantly associated with the age group 2 – 10 years amongst the rural and urban populations. The age group 2-10 years accounted for 21.8% of malaria cases in the entire health district. A study carried out in Yagoua and Maga, in the Far North Region of Cameroon, as far back as 1985, also showed that children between 5 and 9 years old had the highest prevalence of malaria infection [19]. Several studies have shown that parasite prevalence rates in children aged 2–10 years are reliable indicators of malaria endemicity [20, 21]. Based on the classification scheme of malaria endemicity reported elsewhere [22], the infection in rural and urban areas of Maroua III health district can be classified as mesoendemic stratum where transmission is high, under normal rainfall conditions and drops during the dry season.

The distribution of malaria infection in Maroua III health district is heterogeneous and varies greatly between rural and urban settlements within the population. In rural settlements, malaria infection was found to decrease with an increasing level of education, with those who have not gone to school bearing the highest burden (58.9%). Likewise, in urban settlements, malaria infection was found to decrease with levels of education and the infection was significantly associated with levels of primary and nursery education levels. This may reflect the inadequate knowledge and poor practices of preventive strategies against malaria. Based on occupation, malaria was significantly associated with pupils and students in the entire health district. In fact, pupils and students accounted for more than one-third (36.8%) of the infected population in Maroua III health district. This implies that periodical screening and treatment of pupils and students in schools, during the peak malaria season, can decrease the burden of the disease by a third.

Bivariate analysis revealed that malaria was significantly associated with occupants of houses with mud walls, roofed with aluminum sheets or grass in the entire health district, when compared with occupants of houses built with blocks and roofed with aluminum sheets. This could result from the practice that the eaves of houses made with mud are not usually covered, while those of houses made of cement blocks and roofed with aluminum sheets are usually covered, to make the building more embellished. This prevents the entry of mosquitoes into cement block buildings unlike in mud houses, which agrees with the fact that open eaves are known to permit entry of mosquitoes into buildings [23]. Most of the houses in rural areas are constructed with mud/grass or aluminum roofs, compared to urban settings where houses are constructed with cement/aluminum sheet roofs.

Bivariate analysis also showed that occupants of houses without ceilings were significantly associated with malaria infection. The absence of a ceiling permits free movement of mosquitoes into the house, through the eaves. This is in agreement with studies carried out in East and West Africa, where open eaves and absence of ceilings in houses has been associated with increased mosquito nuisance and higher levels of malaria infection, compared to occupants of houses with ceilings and closed eaves [23, 24, 25]. Likewise, from bivariate analysis, malaria was significantly associated with users of pit toilets, over users of water system toilets, in the entire health district. Pit toilets may serve as breeding sites for mosquitoes, which are released through the open mouth of the pit toilet, unlike water system toilets which have enclosed septic tanks from which mosquitoes cannot escape. These findings suggest that the presence of standard houses is beneficial to the occupants and the surrounding community by reducing the risk of malaria infection.

Long-lasting insecticidal nets (LLINs) are one of the main vector control strategies recommended by the World Health Organization for the control and elimination of malaria, but coverage continues to be moderate in many parts of sub-Saharan Africa. However, our study showed a high usage of 90.4% in rural settlements and 84% in urban settlements. Malaria was not significantly associated with non-users of LLINs although the prevalence was higher amongst this group, when compared to users of LLINs. This high coverage of LLINs in Maroua III HD could be due to the presence of more than two household members in almost all households and frequent distribution of LLINs to pregnant women during anti-natal clinics. Previous studies have shown that households whose family sizes are >2 have more chances of owning and using LLINs compared to their counterparts living in households with family size ≤ 2 [38,39]. Even though usage of LLINs was not significantly associated with malaria infection, it shows that sleeping under LLINs is good, but screening of windows with mosquito nets is more beneficial, in this study area. This is in agreement with findings in Yaounde, Cameroon, where screens on Windows were significantly associated with fewer mosquitoes collected indoors [26].

Also, our findings revealed that age of LLINs was associated with malaria infection, whereby households using LLINs older than three years are more prone to malaria in both rural and urban communities of the district. This may be due to the fact the nets may have lost their efficacy and become damaged. This suggests that not retreating old bed nets, because they are LLINs, may have increased the risk of malaria in these localities.

The presence of crops around homes was significantly associated with its occupants being infected with malaria. This corroborates with observations in Bolifamba, located in the South West Region of Cameroon, where malaria was significantly associated with the presence of bushes around homes [27]. Good hygiene is universally known as one of the most efficacious and straightforward measures to prevent disease transmission [40]. In the rural areas of the district, the source of drinking water was found to be associated with malaria infection, with households who obtained their water from open wells more prone to malaria infection. This finding indicated that incremental improvements to water sources in villages across the rural settlements in the District might be considered a potential intervention for the prevention and control of malaria in the long term because water-associated vector-borne diseases (including malaria and many neglected tropical diseases) continue to be a major public health problem in many developing countries [41].

In both rural and urban settlements, it was also observed that geometric mean parasite density decreased with increase in age, which is indicative of the acquisition of naturally acquired immunity to malaria, following repeated infection over the years [28]. The practice of living in the same house with domestic animals, such as goats and cows, was not associated with protection from malaria. This is contrary to the observed 27.2% of blood fed mosquitoes captured in the neighboring region, with similar conditions, were composed of sheep and cow blood <u>[29]</u>. These animals could have served as an alternative source of blood meal, to prevent mosquitoes from going after human blood.

The effectiveness of insecticide-based malaria vector control interventions in Africa is threatened by the spread and intensification of pyrethroid resistance in targeted mosquito populations. These results suggest the possibility of widespread resistance of mosquitoes to permethrin and deltamethrin throughout Maroua III Health District. Les campagnes experimentales d’eradication du paludisme dans le Nord de la Republique du Cameroun also reported mosquitoes’ resistance to pyrethroids in the 1950s [30]. A review of the evolution of insecticide resistance to the malaria vectors in Cameroon from 1990 to 2017 reported an increase in the mosquito’s population resistance to insecticides due to an increased use of treated bed nets, insecticide sprays and the use of insecticides in agriculture [31] and this showed that insecticide resistance can be recognized as a serious threat for control interventions, implemented to fight against malaria. On the whole, mosquitoes in Maroua III Health District showed higher resistance to permethrin than deltamethrin, with values of 83.85% and 93.57% percent susceptibility, respectively. It was also observed that the mortality rate of *Anopheles* in urban areas of the district was higher (95.83%) as compared to mortality in rural areas (91.39%) for the insecticide deltamethrin. This may be due to the high use of insecticides in agriculture within rural areas, which can promote resistance. Studies conducted on *Anopheles gambiae* distribution and insecticide resistance in Douala and Yaoundé showed a high prevalence of insecticide resistance in mosquitoes originating from agricultural-cultivated sites compared to other sites [32]. Also, resistance was higher against permethrin as compared to deltamethrin, suggesting deltamethrin may be more effective than permethrin. This may be due to the fact that the first LLINs distributed in Cameroon before 2016 were impregnated with permethrin and consequently, mosquitoes may have developed resistance to this insecticide over time. A higher mortality of *Anopheles coluzzii* from deltamethrin than permethrin has been reported in the Guinea savanna of Cameroon [33], and a similar trend of greater resistance to permethrin over deltamethrin has also been reported in northern Benin [34]. Although studies on multiple insecticide resistance mechanisms in *Anopheles gambiae* populations from Cameroon showed that the *Anopheles arabiensis* population sampled in Pitoa health area were more susceptible to permethrin than deltamethrin [35].

## CONCLUSION

We observed different variables associated with malaria between rural and urban settlements of Maroua III HD, although there are common risk factors shared across areas. These findings suggest that some interventions such as the use of new mosquito bed nets may help to reduce malaria risk among the population. People in rural areas may potentially benefit in having well-constructed/protected wells for their source of water, whilst people living in urban areas should be encouraged not to farm around their houses to reduced mosquito’s habitation sites. The usage of LLINs older than 3 years and the presence of crops around homes are significant risk factors for malaria infection, and school children bear the greatest burden of the infection. Deltamethrin is a better alternative for impregnation of insecticide treated bed nets than permethrin.

## Data Availability

The data generated from this study are available from the corresponding author on reasonable request.

## ACKNOWLEDGEMENT

The authors are grateful to CDC, 1600 Clifton Road. NE, Atlanta, GA, USA for providing the CDC Bottle Bioassay test kits. We also thank Mr. Yakubou and Mr. Akim Mechewere who assisted in questionnaire administration and facilitated communication in Fufulde language, as well as Miss Kika Delphine who translated the questionnaire from English to French. Special thanks to our Dr. Ngoran, an entomologist for identification of Anopheles species and to all our parasitologists at the University of Buea Laboratory for Emerging Infectious Diseases.

## ABBREVIATIONS

LLINs: long-lasting insecticidal Nets
HD: health district

## AUTHORS’ CONTRIBUTIONS

RBN: conceived the topic. RBN and SFN: wrote the proposal, while SFN: Took part in data collection. RBN, SFN, SNE VPKT: took part in data analysis and interpretation. RBN and SFN: wrote the first draft of the manuscript and all the authors read and approved of the final the manuscript.

## ETHICS APPROVAL AND CONSENT TO PARTICIPATE

Participants were informed on the potential benefit and aim of the study before obtaining their written consent. Written informed consent was obtained from the parent/guardian of each participant under 18 years of age. Parents or guardians gave assent for minors by filling out and signing the assent form. Ethical approval was obtained from the Faculty of Health Science Institutional Review Board of the University of Buea reference number: 2019/979-05/UB/SG/IRB/FHS. Administrative authorization was obtained from the Far North Regional Delegation of Public Health reference number: 374/ar/19/MINSANTE/SG/DRSP/EN/YT/MRA. Subsequent authorizations were sought from village heads (Lawanats), quarter heads and community leaders of concerned localities.

## CONSENT FOR PUBLICATION

Not applicable

## Legend for Supporting Information

**S1 File:** Data capture questionnaire English

**S2 File:** Data capture questionnaire French

